# RNase HII-assisted amplification (RHAM) for rapid point-of-care monkeypox detection

**DOI:** 10.64898/2026.04.01.26349928

**Authors:** Julian Kamhieh-Milz, Sundrela Kamhieh-Milz, Franziska Schwarz, Janine Michel, Andreas Nitsche, Andreas Puyskens

## Abstract

Mpox poses an ongoing global public health threat, with case numbers rising beyond traditionally endemic regions in Central and Western Africa. Rapid detection of the causative agent, the Monkeypox virus (MPXV), is critical for outbreak control, yet laboratory infrastructure and trained personnel remain scarce in many affected areas. Point-of-care molecular diagnostics offer a practical solution by enabling timely testing without specialized equipment or elaborate nucleic acid extraction. We evaluated the performance of an extraction-free RNase HII-assisted amplification (RHAM) assay for MPXV detection by Pluslife Biotech, a novel isothermal amplification technology providing results in under 30 minutes. The Pluslife RHAM test demonstrated pan-MPXV clade reactivity, detecting all four MPXV clades (Ia, Ib, IIa, IIb) with high analytical sensitivity and no cross-reactivity to other poxviruses or other clinically relevant pathogens. The assay proved compatible with diverse clinical specimen types, including lesion swabs, oropharyngeal swabs, rectal swabs, urine, semen, and wound exudate. As part of routine diagnostics at the German Consultant Laboratory for Poxviruses, in a comprehensive evaluation of 206 clinical specimens against diagnostic real-time PCR, the Pluslife RHAM test achieved a diagnostic sensitivity of 94.2% (95% CI: 85.8–98.4%) and a specificity of 100% (95% CI: 97.3–100%). Notably, samples with higher viral loads (Ct <30) showed 100% sensitivity. Time-to-result correlated significantly with viral load, enabling faster diagnosis in high-viral-load cases. The Pluslife RHAM test represents a practical, sensitive, and rapid point-of-care solution for MPXV detection in resource-limited settings, combining strong analytical performance with operational simplicity to support timely outbreak response and clinical decision-making.

## Introduction

Monkeypox virus (MPXV) is a double-stranded DNA virus of the genus *Orthopoxvirus* (OPXV) within the family *Poxviridae*, and is closely related to variola virus, the causative agent of smallpox (1). There are two distinct MPXV clades: clade I (with subclades Ia and Ib) and clade II (with subclades IIa and IIb) (2). First isolated in 1958 from captive monkeys (3), MPXV is transmitted zoonotically through close contact with infected animals, particularly rodents and small mammals, and increasingly through direct human-to-human transmission (4). The World Health Organization (WHO) has declared mpox a Public Health Emergency of International Concern (PHEIC) twice: In 2022, due to the global spread of clade IIb, and in 2024 following the emergence and spread of clade Ib in the Democratic Republic of Congo (DRC) and neighbouring countries (2). The outbreak has persisted into 2025, extending beyond traditionally endemic regions in Central and West Africa to affect multiple countries worldwide (5). Ongoing transmission, evolving clinical presentations, and challenges in vaccine distribution have continued to raise global public health concerns.

Effective outbreak containment relies on prompt access to accurate, locally deployable diagnostic testing. Real-time PCR remains the gold standard for MPXV detection, offering species-level differentiation of OPXV and clade-level identification of MPXV. However, PCR typically requires a specialized laboratory infrastructure and trained laboratory personnel, which are frequently unavailable in resource-limited settings, including many regions where mpox is endemic.

Point-of-care (POC) diagnostics have demonstrated substantial value in resource-limited settings, improving patient outcomes and enabling cost-effective management of infectious diseases such as malaria, AIDS and COVID-19. Lateral flow assays (LFAs) became widely used in both professional and self-test settings during the COVID-19 pandemic, for their ease-of-use and rapid turnaround. However, MPXV LFAs evaluated to date did not meet the WHO sensitivity criteria, and their routine application is currently not recommended (6,7). In contrast, molecular point-of-care tests for MPXV have demonstrated substantially improved performance compared to antigen LFAs, with the Cepheid Xpert Mpox and SD Biosensor STANDARD M10 MPX/OPX achieving 74–79% overall sensitivity using lesion samples in independent multi-site evaluations (7). However, none of the tested molecular point-of-care tests met the WHO target product profiles requirements for mpox (8) when evaluated with prospective samples from endemic settings.

Molecular nucleic-acid based rapid detection methods offer a promising alternative to laboratory-based PCR diagnostics and antigen LFAs. In particular, loop-mediated isothermal amplification (LAMP) has emerged as an attractive alternative for use in resource-limited settings, combining high sensitivity with rapid turnaround times while eliminating the need for thermal cyclers and extensive laboratory infrastructure.

RNase HII-assisted amplification (RHAM) represents an advanced isothermal amplification technology that integrates LAMP-based amplification with an RNase HII-dependent reporter system. The RHAM platform has previously demonstrated sensitivity comparable to laboratory-based real-time PCR for SARS-CoV-2 detection (9) while delivering results within 7–35 minutes. Here, we evaluate the analytical and clinical performance of the Pluslife RHAM test for MPXV detection across all clades, assess its compatibility with diverse clinical specimen types, and compare its diagnostic accuracy against routine real-time PCR as part of the diagnostics in the German Consultant Laboratory for Poxviruses.

## Materials and Methods

### Specimens used for performance evaluation

Both virus culture supernatants and clinical specimens were used for comparative performance evaluation of the Pluslife RHAM test. MPXV cultures were performed on VeroE6 cells (#85020206, ECACC) as previously described (10). Culture supernatants of MPXV clades Ia, IIa, and IIb were heat-inactivated and γ-irradiated prior to use. MPXV clade Ib culture supernatants and MPXV-positive clinical specimens were used as native material and handled under BSL-3 conditions. Clinical specimens were collected as part of routine diagnostics at the German Consultant Laboratory for Poxviruses in ZBS 1 at the Robert Koch Institute in Berlin, Germany, between 2022 and 2025. Specimens were either tested directly within 24 hours of laboratory receipt or stored at −70°C until further analysis. The use of clinical specimens to improve MPXV diagnostics obtained ethical approval by the Berliner Ärztekammer (#ETH 44/22).

## Reference diagnostics

### Nucleic acid extraction for real-time PCR and digital PCR

Nucleic acid extraction for real-time PCR was performed using the QIAamp DNA Blood Kit (QIAGEN, Hilden, Germany). Prior to extraction, samples were diluted 1:2 by adding 100 µL of sample to 100 µL 1 x phosphate buffered saline (PBS) to maintain consistent nucleic acid concentrations before and after extraction following the protocol of the QIAamp DNA Blood Kit. Hence, a total volume of 200 µL diluted sample was extracted and eluted in 100 µL nuclease-free H_2_O. Extractions were performed using a QIAcube Connect (QIAGEN) automated extractor. Extracted DNA was either stored at 4°C for up to 2 days or at −70°C for long-term storage.

### Real-time PCR and digital PCR analysis

Confirmation of sample positivity and MPXV clade-typing was performed using real-time polymerase chain reaction (PCR) as previously described (10) as part of routine diagnostics at the German Consultant Laboratory for Poxviruses in ZBS 1, Robert Koch Institute (accredited according to DIN ISO 15189). For absolute quantification of MPXV viral copy numbers, digital PCR (dPCR) was performed using primers and probe targeting the MPXV generic G2R gene previously described (10). dPCR was performed with the QIAcuity Probe PCR Kit according to the manufacturer’s instructions on a QIAcuity One 5-Plex instrument (QIAGEN). A volume of 5 µL template was added to each real-time and dPCR reaction. Viral concentrations are shown as viral copies (cp)/PCR reaction.

### Pluslife RHAM test evaluation

Performance evaluations were conducted using the Pluslife Monkeypox Virus Nucleic Acid Test (Guangzhou Pluslife Biotech Co., Ltd., Guangdong, China) (11) on the Pluslife Mini Dock isothermal amplification and reading device (Figure 1). Instead of swabs, 50 µl of sample dilutions or transport medium were added directly to the test buffer, and subsequent testing was performed in accordance with manufacturer’s instructions.

**Figure 1.**
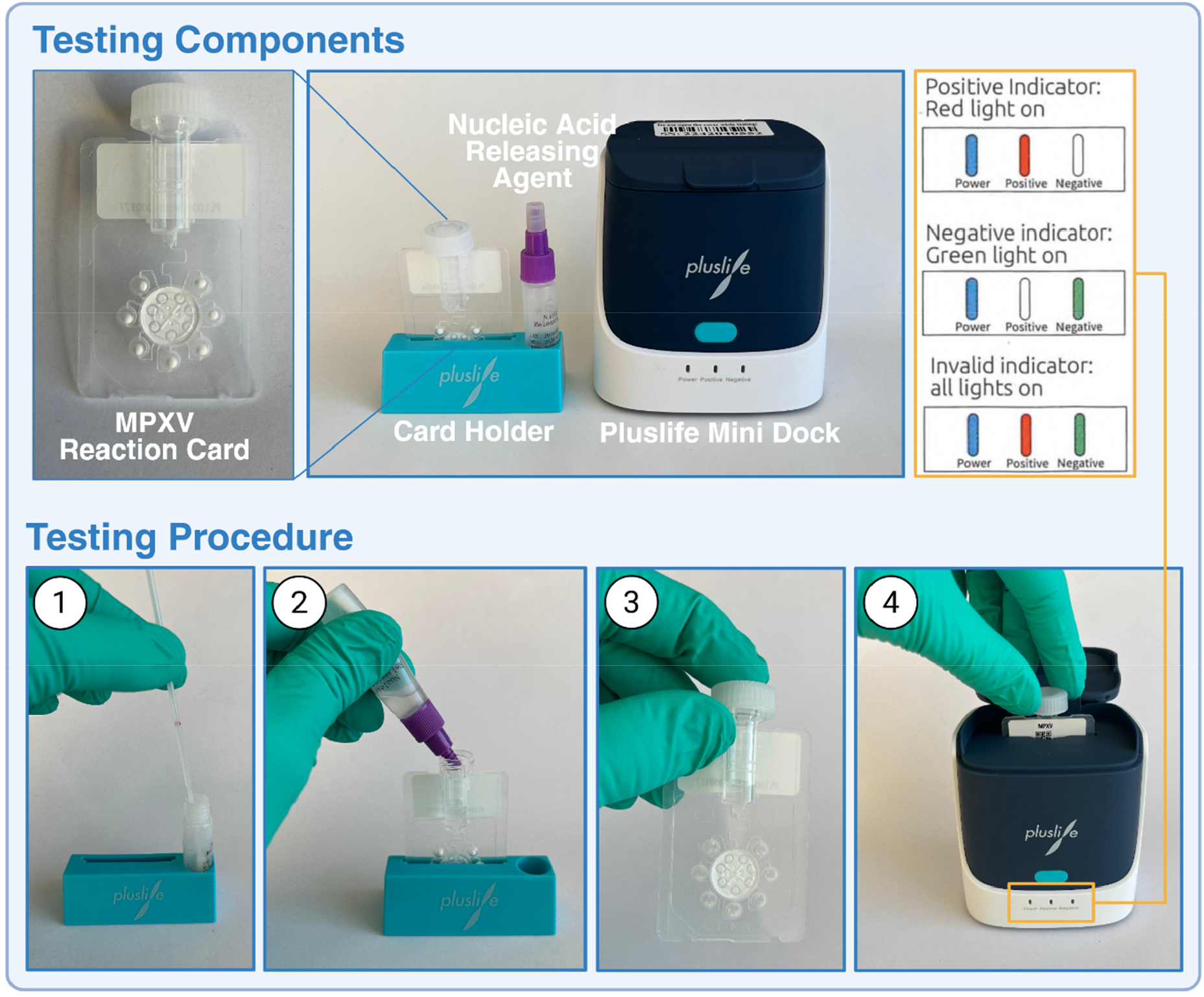
Pluslife RHAM MPXV components and testing procedure. Pluslife RHAM testing components: MPXV Reaction Card containing lyophilized testing reagents, card holder for sample preparation, Nucleic Acid Releasing Agent 02 for extraction-free sample processing, and Pluslife Mini Dock reader device. Testing procedure: (1) specimen is collected using a swab and inserted to the vial containing the Nucleic Acid Releasing Agent; (2) Sample containing Nucleic Acid Releasing Agent is added to the Reaction Card via the provided dripper cap; (3) Pressing the cap on the top of the Reaction Card transfers sample containing Releasing Agent into reaction chambers of the Reaction Card; (4) Lastly, the Reaction Card is inserted into the Mini Dock for isothermal amplification and subsequent result readout. Result interpretation is indicated by LED lights: red (positive), green (negative), or all lights illuminated (invalid).

### Analytical specificity and sensitivity evaluation

Test specificity was assessed using a panel of native culture supernatants from various OPXV species: cowpox virus, vaccinia virus; camelpox virus, and ectromelia virus; non-OPXV poxviruses: Orf virus and molluscum contagiosum virus; as well as other clinically relevant viral pathogens for differential diagnosis: varicella zoster virus, herpes simplex virus-1, and -2. Respective viral loads were determined using pathogen-specific in-house real-time PCR assays according to standard protocols at ZBS 1. To pass the internal control of the Pluslife RHAM test for samples derived from viral cell culture, additionally 2 µL of uninfected VeroE6 cell suspension were spiked to the test buffer directly before analysis. Results were recorded using the Pluslife Pad Desktop-Software (Version SV4.2.1.20240403).

### Pluslife RHAM test performance in routine poxvirus diagnostics

Various clinical specimen types were used for performance evaluation, including lesion swabs, crust, oropharyngeal swab, semen, urine and wound exudate provided either as archived (stored at −70°C) or fresh samples. Crust, lesion, and oropharyngeal swab samples were stored in 1 mL PBS. Prior to testing and real-time PCR analysis, native samples were diluted 1:5 by adding 50 µL of sample to 200 µL PBS. As before, 50 µL of diluted sample were directly added to the respective test buffer, and POC testing was performed according to manufacturer’s instructions. To evaluate the performance of the Pluslife RHAM test in a real-world diagnostic setting, some of the clinical specimen testing was conducted in parallel to the routine real-time PCR poxvirus diagnostic workflow at the German Consultant Laboratory for Poxviruses, from January 2025 to August 2025. The Pluslife RHAM test used 50 µL of viral transport medium (VTM) or lesion swabs resuspended in 1 mL PBS added to the test buffer without prior sample extraction. Finally, results from the Pluslife RHAM test and real-time PCR were compared to determine diagnostic performance.

## Statistical analysis

Diagnostic sensitivity and specificity were calculated from the 2×2 contingency table comparing Pluslife RHAM test results against diagnostic real-time PCR as the reference standard. Exact 95% confidence intervals (CIs) for sensitivity and specificity were calculated using the Clopper-Pearson exact binomial method. The correlation between viral load (Ct values from generic MPXV real-time PCR) and time-to-result from the Pluslife RHAM test was assessed using Spearman’s rank correlation coefficient. A two-tailed p-value of <0.05 was considered statistically significant. Statistical analyses were performed using GraphPad Prism (Version 9.1.0).

## Results

### Validation of the Pluslife RHAM test

#### Cross-reactivity to diagnostically relevant pathogens

To assess the analytical specificity of the Pluslife RHAM test, a panel of cell culture propagated viral pathogens relevant to the differential diagnosis of MPXV was evaluated (Table 1). The Pluslife RHAM test demonstrated high analytical specificity, yielding negative results for all tested non-MPXV pathogens in the specificity panel.

**Table 1.** Specificity of the Pluslife MPXV RHAM test. Testing performed in duplicates. OPXV, Orthopoxvirus; *neg*, negative.

|  | **Virus** | **In-house**<br>**PCR**<br>**[Ct]** | **Result**<br>**Pluslife** |
| --- | --- | --- | --- |
| OPXV | Cowpox virus (Brighton Red) | 17.6 | *neg* \| *neg* |
|  | Vaccinia virus | 15.2 | *neg* \| *neg* |
|  | Camelpox virus | 19.0 | *neg* \| *neg* |
|  | Ectromelia virus | 15.9 | *neg* \| *neg* |
| Non-OPXV<br>viruses relevant<br>for differential<br>diagnosis | Molluscum contagiosum virus | 21.0 | *neg* \| *neg* |
|  | ORF virus | 15.2 | *neg* \| *neg* |
|  | Varicella Zoster virus | 18.5 | *neg* \| *neg* |
|  | Herpes Simplex virus 1 | 18.7 | *neg* \| *neg* |
|  | Herpes Simplex virus 2 | 18.4 | *neg* \| *neg* |

### Analytical sensitivity of the Pluslife RHAM test

To evaluate the analytical sensitivity of the Pluslife RHAM test, serial dilutions of MPXV culture supernatants of clades Ia, Ib, IIa, IIb were tested in the Pluslife RHAM test and quantified after nucleic acid extraction and subsequent MPXV real-time PCR and dPCR (Table 2). Depicted concentrations refer to virus cp/PCR reaction and therefore should not be directly compared with the Pluslife RHAM test, which uses native, unextracted samples.

**Table 2.** Performance of the Pluslife RHAM test with MPXV clades from cell culture supernatant. Real-time PCR = generic (G2R) MPXV PCR. Grey areas indicate the border of the lower detection limits of the Pluslife RHAM test for each clade. Pos, positive; *neg*, negative; dPCR, digital PCR; Cp, viral copies; Ct, quantification threshold; n.a. = not applied.

| Clade | dPCR<br>[cp/PCR reaction] | real-time PCR<br>[Ct value] | Pluslife RHAM<br>test | Repeat testing |
| --- | --- | --- | --- | --- |
| Ia | 3,723 | 25.2 | pos \| pos | n.a. |
|  | 200 | 29.4 | pos \| pos | pos \| pos \| pos \| pos \| *neg* \| pos |
|  | 16 | 33.1 | *neg* \| *neg* | *neg* \| pos \| *neg* \| *neg* \| pos \| *neg* |
|  | 1 | 37.4 | *neg* \| *neg* | n.a. |
| Ib | 10,869 | 24.3 | pos \| pos | n.a. |
|  | 723 | 28.2 | pos \| pos | n.a. |
|  | 44 | 32.7 | pos \| pos | pos \| *neg* |
|  | 8 | 34.6 | pos \| pos | *neg* \| pos \| pos \| pos \| pos \| pos |
| IIa | 39,090 | 24.2 | pos \| pos | n.a. |
|  | 2,765 | 28.0 | pos \| pos | pos \| pos \| pos \| pos \| pos \| pos |
|  | 226 | 31.7 | pos \| pos | *neg* \| *neg* \| *neg* \| pos \| pos \| *neg* |
|  | 23 | 35.5 | *neg* \| *neg* | pos \| *neg* \| *neg* |
|  | 1 | 37.6 | *neg* \| *neg* | n.a. |
| IIb | 40,356 | 22.1 | pos \| pos | n.a. |
|  | 3522 | 25.7 | pos \| pos | n.a. |
|  | 292 | 29.7 | pos \| pos | pos \| pos \| pos \| pos \| pos \| pos |
|  | 22 | 32.5 | *neg* \| *neg* | *neg* \| *neg* \| *neg* \| *neg* \| *neg* \| *neg* |

For clade Ia and Ib, the Pluslife RHAM test detected 7 of 8 replicates corresponding to 200 and 8 cp/PCR reaction, respectively (Figure 2). MPXV clades IIa and IIb were reliably detected at viral concentrations of 2,765 cp/PCR reaction and 292 cp/PCR reaction, respectively (Figure 2).

**Figure 2.**
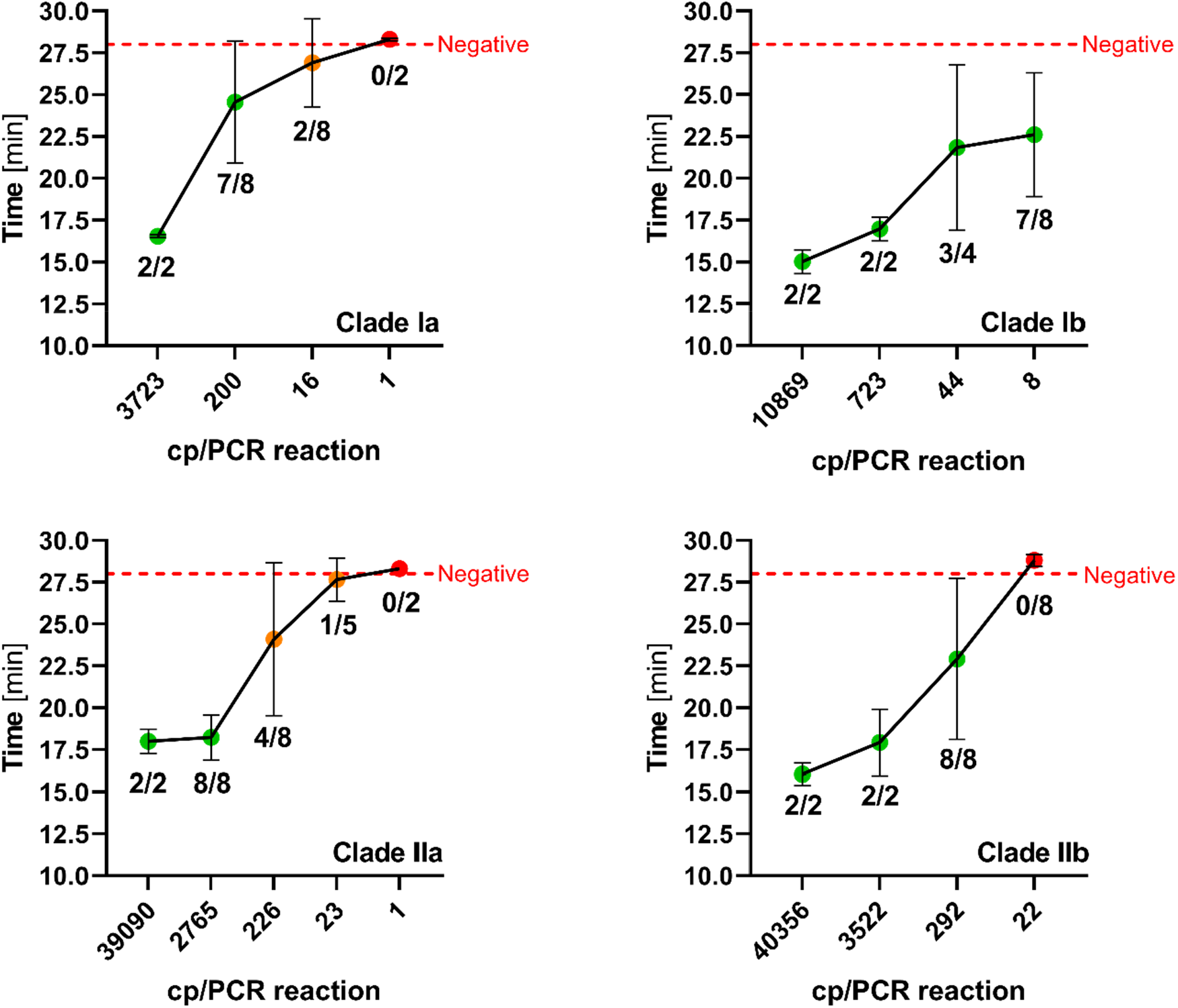
Analytical sensitivity of the Pluslife RHAM test across MPXV clades. Serial dilutions of cell culture-propagated MPXV clades Ia, Ib, IIa, and IIb were tested to determine the limits of detection for each clade. The x-axis shows viral loads in copies per PCR reaction (cp/PCR reaction), and the y-axis shows the time to positivity in minutes in the Pluslife. Circles indicate mean time-to-result with standard deviation. Numbers below each data point indicate the fraction of positive results (positive tests/total tests performed). The red dashed line represents the assay cutoff, above which results are negative.

The Pluslife platform terminates amplification once a predefined signal threshold is reached. We observed a statistically significant positive correlation between time-to-result and viral load (Spearman’s r = 0.7613, p < 0.0001; Figure 3).

**Figure 3.**
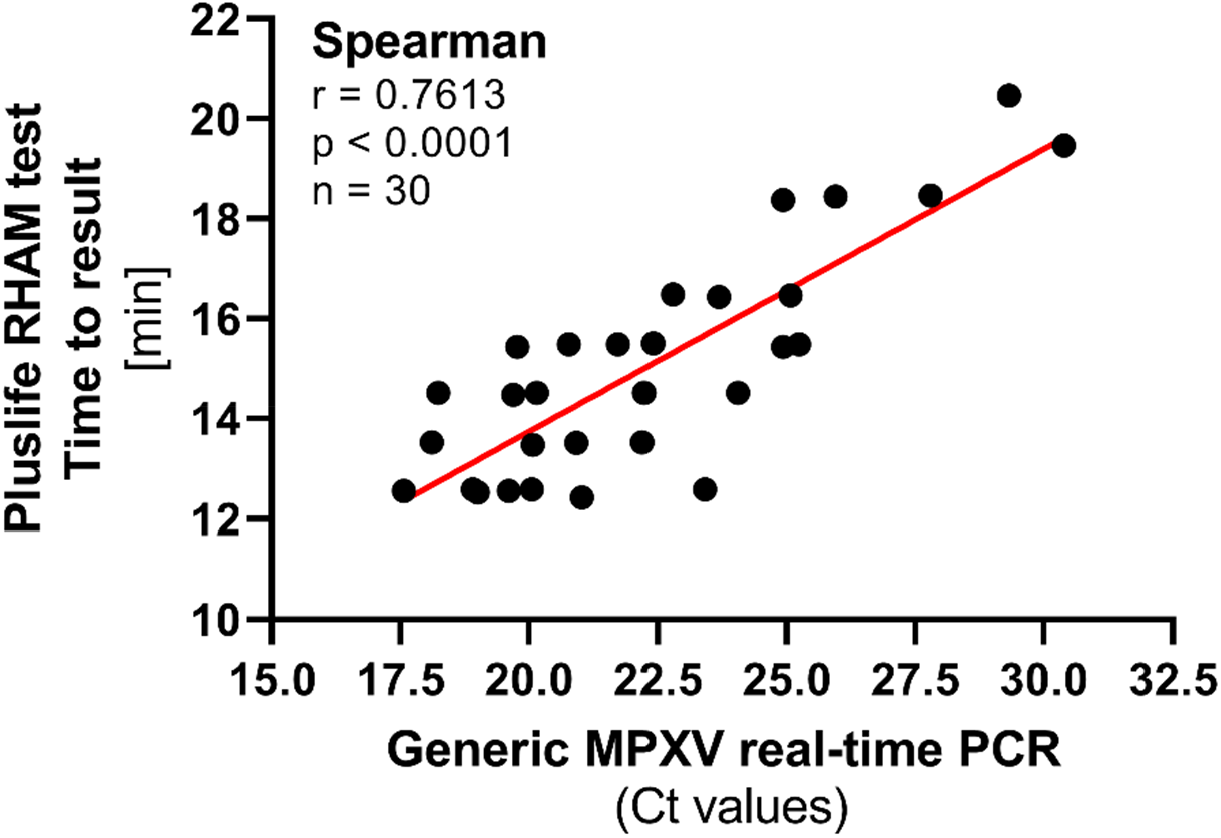
Correlation of viral load and time-to-result as generated by the Pluslife RHAM test. The Pluslife platform terminates the amplification process once a predefined threshold of signal intensity is reached. An analysis of n = 30 MPXV-positive lesion swab clinical specimens demonstrates a statistically significant positive correlation between viral load in a specimen, as indicated by Ct values, and time-to-result (Spearman’s r = 0.7613, p < 0.0001).

### Evaluation of the Pluslife RHAM test using different clinical specimen types

Evaluation across a range of MPXV clade IIb-positive clinical specimen types suggests suitability of the Pluslife RHAM test for diverse sample types (Table 3). Crust (mean Ct: 22.1 ± 4.2), oropharyngeal swabs (mean Ct: 22.5 ± 2.5), and rectal swabs (mean Ct: 20.0 ± 2.8) were reliably detected in all five specimens tested. Wound exudates (mean Ct: 23.9 ± 2.4) and urine samples (mean Ct: 25.8 ± 2.8) were detected in four out of five samples. Of the five semen samples, which overall showed the lowest viral loads (mean Ct: 30.8 ± 4.0), two samples were not detected, one negative and one invalid, both with Ct values >32.5.

**Table 3.** Compatibility of different clinical specimens with the Pluslife RHAM technology. Shown in parentheses are corresponding quantification threshold (Ct) values for each sample as tested in the generic (G2R) MPXV real-time PCR. All specimens contain MPXV clade IIb. n, number of samples tested; pos, positive; *neg*, negative.

| Sample type | n | Pluslife RHAM test results (Ct generic MPXV real-time PCR) |  |  |  |  |  |
| --- | --- | --- | --- | --- | --- | --- | --- |
| Crust | 5 | pos<br>(18.3) | pos<br>(19.1) | pos<br>(19.9) | pos<br>(26.1) | pos<br>(27.1) | **5 / 5**<br>**(22.1 ± 4.2)** |
| Oropharyngeal<br>swab | 5 | pos<br>(19.6) | pos<br>(20.7) | pos<br>(22.5) | pos<br>(23.7) | pos<br>(26.1) | **5 / 5**<br>**(22.5 ± 2.5)** |
| Rectal swab | 5 | pos<br>(16.1) | pos<br>(19.3) | pos<br>(19.8) | pos<br>(21.2) | pos<br>(23.6) | **5 / 5**<br>**(20.0 ± 2.8)** |
| Wound exudate | 5 | pos<br>(21.1) | pos<br>(22.2) | pos<br>(23.8) | pos<br>(25.0) | *invalid*<br>(27.3) | **4 / 5**<br>**(23.9 ± 2.4)** |
| Urine | 5 | pos<br>(21.6) | pos<br>(24.2) | *invalid*<br>(27.1) | pos<br>(28.0) | pos<br>(28.1) | **4 / 5**<br>**(25.8 ± 2.8)** |
| Semen | 5 | pos<br>(25.0) | pos<br>(28.3) | pos<br>(32.3) | *neg*<br>(33.9) | *invalid*<br>(34.3) | **3 / 5**<br>**(30.8 ± 4.0)** |

### Diagnostic sensitivity and specificity of the Pluslife RHAM test

A total of 206 clinical lesion swab samples (69 MPXV PCR-positive, 137 MPXV PCR-negative) collected as part of routine diagnostics at the German Consultant Laboratory for Poxviruses in ZBS 1 at the Robert Koch Institute, were analysed to evaluate the diagnostic sensitivity and specificity of the Pluslife RHAM test (Table 4). Out of 69 PCR-positive specimens tested, the Pluslife RHAM test falsely reported four specimens as negative. PCR-negative specimens were all detected correctly as MPXV-negative by the Pluslife RHAM test. Accordingly, the Pluslife RHAM test demonstrated a diagnostic sensitivity of 94.2% (95% CI: 85.8–98.4%) and a specificity of 100% (95% CI: 97.3–100%).

**Table 4.** Clinical performance evaluation. Total number of true positive samples (PCR-positive) = 69; Total number of true negative (PCR-negative) = 137; Pluslife RHAM sensitivity = 94.2% (95% CI: 85.8–98.4%); specificity = 100% (95% CI: 97.3–100%). Confidence intervals calculated using the Clopper-Pearson exact binomial method.

|  |  | Pluslife RHAM test |  |
| --- | --- | --- | --- |
|  |  | positive | negative |
| Dx real-time PCR | positive | **65** | **4** |
|  | negative | **0** | **137** |

## Discussion

In remote and underserved regions, the absence of laboratory infrastructure frequently limits access to timely diagnostic services. POC testing plays a critical role in outbreak control by enabling rapid identification and isolation of infected individuals. In the past, POC diagnostics have proven successful for detecting pathogens including HIV, SARS-CoV-2, dengue virus, malaria parasites, and other tropical diseases in resource-limited settings (12–15). While antigen-based rapid tests offer advantages, such as ease-of-use and minimal equipment requirements, nucleic acid-based POC platforms can overcome key limitations of antigen detection, including reduced sensitivity and specificity. LAMP has already demonstrated field utility across diverse settings (16), and a LAMP-based assay, targeting pulmonary tuberculosis, has recently received WHO approval as alternative to real-time PCR (17). RHAM represents a recent innovation: The use of recombinase proteins in RHAM enables probe-based detection and may therefore further improve sensitivity and specificity (18).

Test specificity is critical for accurate and differential diagnosis, particularly given clinical overlap between MPXV and other infections with similar clinical presentation. In our study, the Pluslife RHAM test displayed complete specificity for MPXV, with no cross-reactivity observed against other Orthopoxviruses, Parapoxviruses, Molluscum contagiosum, and other viruses relevant for differential diagnoses (VZV, HSV-1, HSV-2).

In our comprehensive study of 206 clinical specimens (69 MPXV PCR-positive and 137 MPXV PCR-negative), the Pluslife RHAM test achieved an overall sensitivity of 94.2% (95% CI: 85.8– 98.4%) and specificity of 100% (95% CI: 97.3–100%). Notably, the four MPXV PCR-positive specimens that were not detected by the Pluslife RHAM test had Ct values >30 and were archived (previously frozen and stored) specimens. These results are consistent with findings from Cavuto et al., who clinically validated a LAMP-based assay on 164 samples, including 51 MPXV-positive samples, and reported a sensitivity of 96.1% and specificity of 100% for Orthopoxviruses, and a sensitivity of 94.1% and specificity of 100% for MPXV (19).

The ability of the Pluslife RHAM test to detect all MPXV clades and successful handling of different specimen types enhances its applicability across diverse outbreak scenarios. Specimen types can influence assay performance, with the most consistent results obtained from lesion swabs, crusts, oropharyngeal swabs and rectal swabs — sample types that also display high viral loads in line with previous studies and clinical recommendations (20–22). These sample types are readily accessible from symptomatic individuals and can be easily collected in outpatient or emergency settings, making them well-suited for rapid POC workflows. Detection in wound exudate, semen and urine was also demonstrated using the Pluslife RHAM test, though with greater variability, suggesting these sample types may serve better as supplementary specimens rather than primary diagnostic samples (20,23). Semen has been shown to frequently inhibit PCR reactions (24,25), which indicates that this sample type might also be challenging for extraction-free RHAM/LAMP reactions, potentially explaining the inferior performance of the Pluslife RHAM test with this specimen type.

The Pluslife RHAM platform terminates amplification once a predefined signal threshold is reached, enabling faster results for specimens with high viral loads. While a previous evaluation using the Pluslife RHAM platform for SARS-CoV-2 testing did not observe a relationship between viral load and time-to-result (9), in our study time-to-result correlated significantly with viral load. This correlation suggests that time-to-result may serve as a semi-quantitative indicator of viral load, potentially offering additional clinical utility for future specimen triage and interpretation.

Rapid diagnostic tests improve outcomes by enabling earlier treatment initiation and isolation, particularly for infectious diseases with high transmission rates (26,27). For MPXV, rapid diagnosis facilitates timely infection control measures, contact tracing, and appropriate clinical management. Cost-benefit analysis for COVID-19 antigen-testing in Germany demonstrates that POC testing can accelerate clinical decision-making, as results were typically available one day earlier than routinely used laboratory-based PCR tests (28). The Pluslife RHAM platform addresses key barriers to molecular diagnostics in resource-limited settings. Unlike traditional PCR-based diagnostics requiring specialized equipment, stable power supply, cold chain reagent storage, and trained laboratory personnel, the Pluslife platform operates with minimal infrastructure and can be deployed in decentralized settings. The extraction-free workflow further simplifies implementation by eliminating time-consuming sample preparation steps and reducing biosafety requirements. Operator training and proficiency significantly influence POC testing reliability, with user variability representing a recognized source of error in field settings (29). The Pluslife RHAM platform mitigates this challenge through automated result capture, objective threshold-based interpretation, and integrated data logging. Unlike colorimetric or visual read-out systems that require subjective interpretation, the Pluslife system provides standardized, digital results. This design reduces operator-dependent variability while supporting quality assurance and regulatory compliance. However, as with all POC workflows, maintaining diagnostic quality requires robust quality control and quality assurance programs, operator training, and adherence to standardized testing protocols.

Cost-effectiveness remains a critical determinant of diagnostic accessibility in resource-limited settings. More than 11,000 GeneXpert instruments have been deployed across low- and middle-income countries, primarily for tuberculosis diagnosis (30). While this existing infrastructure could theoretically support mpox testing, significant economic barriers persist. A standard 4-module GeneXpert system costs approximately US$17,000 (concessional pricing) with additional annual maintenance costs of US$2,000-3,000 (30). GeneXpert mpox test cartridges are currently priced at approximately US$20 per test, substantially higher than the tuberculosis cartridge price of US$7.97 (31). For resource-constrained settings managing dual burdens of tuberculosis and emerging infectious diseases, such as mpox, these cumulative costs significantly limit testing scale-up. At the time of writing, the Pluslife Mini Dock device can be purchased in Germany for US$298, with a per-test cost for mpox testing of US$11.12 (32,33). Reduced capital requirements and lower per-test costs may enable broader diagnostic access, particularly important for decentralized testing strategies in underserved regions.

## Conclusion and Outlook

The Pluslife RHAM test for MPXV detection demonstrates strong analytical performance across MPXV clades, robust diagnostic accuracy with various clinical specimens and high specificity. The extraction-free workflow, rapid turnaround time (<30 minutes), and minimal infrastructure requirements position this platform as a practical solution for MPXV diagnosis in resource-limited and decentralized settings. Future field evaluation in endemic regions would provide valuable insights into operational feasibility, user acceptability, and public health impact under real-world resource constraints, particularly in settings where laboratory infrastructure is limited or absent.

## Acknowledgements

We thank Guangzhou Pluslife Biotech Co., Ltd., for providing MPXV tests within the scope of this study. We thank Laura Herrmann, Vanessa Riehl, Akin Sesver and Thomas Rinner for technical support.

## Author Contributions

Conceptualization: JKM, SKM, AN, JM, AP

Methodology: JKM, SKM, AN, AP

Investigation: SKM, FS, AP

Formal analysis: JKM, SKM, AP

Data curation: JKM, SKM, AP

Writing – original draft: JKM, SKM, AN, AP

Writing – review & editing: JKM, SKM, AN, JM, FS, AP

Supervision: AN, JM, AP

## Data Availability

All data supporting the findings of this study are available within the manuscript and its tables. Additional data are available from the corresponding author upon reasonable request.

